# The Socioeconomic Impact of COVID-19 in Urban Informal Settlements

**DOI:** 10.1101/2021.01.16.21249935

**Authors:** Diego Gil, Patricio Domínguez, Eduardo A. Undurraga, Eduardo Valenzuela

**Affiliations:** Escuela de Gobierno, Pontificia Universidad Católica de Chile, Santiago, RM, Chile; Department of Research, Inter-American Development Bank, Washington DC; Millennium Nucleus for the Study of the Life Course and Vulnerability Santiago, RM, Chile; Research Center for Integrated Disaster Risk Management (CIGIDEN), Santiago, RM, Chile; Department of Sociology, Pontificia Universidad Católica de Chile, Santiago, RM, Chile

**Keywords:** COVID-19, informal settlements, poverty, employment, spatial inequalities

## Abstract

The COVID-19 pandemic has reached almost every corner of the world. Without a pharmaceutical solution, governments have been forced to implement regulations and public policies to control social behavior and prevent the spread of the virus. There is dramatic evidence of the social and economic effects of these measures and their disparate impact on vulnerable communities. Individuals living in urban informal settlements are in a structurally disadvantaged position to cope with a health crisis such as the COVID-19 pandemic. This paper examines the socioeconomic impact of the crisis brought by the pandemic in informal settlements in Chile. We use a three-wave panel study to compare the situation in informal settlements before and during the health crisis. We show that households living in informal settlements are paying a high toll. Their employment loss is dramatic, substantially larger than the loss reported in the general population, and has particularly affected the inmigrant population. We also find that the pandemic has triggered neighborhood cooperation within the settlements. Targeted government assistance programs have reached these communities; however, this group’s coverage is not enough to counteract the magnitude of the crisis. Our results suggest that governments, the non-profit sector, and the community need to urgently provide economic support and protections to individuals living in informal settlements and consider this opportunity for long-term improvements in these marginalized communities.

**Highlights:**

- Governments have implemented large-scale non-pharmaceutical interventions to control the spread of the COVID-19 pandemic
- These measures have had dramatic social and economic effects on the population, particularly affecting vulnerable communities
- Individuals living in urban informal settlements are in a structurally disadvantaged position to cope with this crisis
- Using panel data, we document a dramatic employment loss among informal settlements dwellers, substantially larger than the general population
- The pandemic has also triggered neighborhood cooperation within the settlements as well as targeted government assistance, but not enough to counteract the magnitude of the economic loss

## Introduction

Since early 2020, the world is experiencing a major health and social crisis caused by the COVID-19 pandemic [1]. As of November 5, 2020, more than 50 million cases and 1.2 million deaths have been reported worldwide [2]. Without a readily available pharmaceutical solution to the COVID-19 crisis, governments globally have implemented regulations and guidelines to control social behavior and prevent the spread of the virus [3-5]. Already there is dramatic evidence of the social and economic effects of the pandemic and the governments’ control measures on the general population [6-12]. Moreover, the pandemic does not affect every social group in the same way. Studies suggest that the health crisis has amplified existing disadvantages along racial, ethnic, gender, and socioeconomic lines [13-18].

Individuals living in informal settlements (often called “slums”) are in a structurally disadvantaged position to cope with a health crisis such as the COVID-19 pandemic [19-23]. Around one billion people live in an informal settlement in the global south, a number that has massively increased in the past decades [24-26]. Informal settlements are heterogeneous places [27]. Their emergence and persistence respond to different social, economic, and even political dynamics of the cities that host these communities. [28-30]. They share some common characteristics, however. Informal settlements typically host vulnerable populations in precarious housing conditions, without tenure security, with insufficient access to basic services [31].

The labor, urban, and social conditions typically present in informal settlements may cause its residents to be disproportionally affected during a health crisis, such as the COVID-19 pandemic. This crisis has the potential to exacerbate the spatial inequalities represented by these communities. For instance, governments in many countries have imposed strict lock-down and stay-at-home orders [3,4,32,33]. However, many individuals in informal settlements have informal, precarious jobs, irregular income, no savings, and need to go out to earn the income they need daily [34-36]. Most governments have also made individual-level behavioral recommendations, including keeping physical distance, washing hands frequently, self-quarantines, and sanitizing surfaces. Many of these regulations assume people live in adequate housing conditions, with sufficient living space and access to basic services, such as water and sanitation, which is not the case in most, if not all, informal settlements [31,37]. Residents of informal settlements often develop strong social networks to share social and economic resources, and fight for their access to public and private services [38,39]. During a health crisis, such as the COVID-19 pandemic, however, the possibilities for social interaction and collaboration are significantly reduced.

In this paper, we examine the socioeconomic impact of the COVID-19 pandemic in informal settlements in Chile. The Ministry of Health reported the first COVID-19 case in the country on March 3rd, and several epidemic control measures quickly followed [40,41]. Less than three weeks following the first reported case, the government had announced the closure of schools and universities, national night curfews, border controls, closure of non-essential businesses, and a strategy of localized lockdowns starting March 22 in seven municipalities. In mid-May, Greater Santiago (∼40% of the Chilean population) was put under lockdown, and by mid-June, half the population in Chile was under lockdown [40]. Research suggests that these mitigation strategies significantly slowed down epidemic growth [42] but generated a large drop in mobility and economic activity [6,43]. The reduction in economic activity does not affect all social groups in the same way, which calls for research on the impact on vulnerable populations such as those in informal settlements.

We base our analysis on a three-wave panel (longitudinal) study of households living in Chile’s informal settlements. The baseline was conducted in person in August 2019 (n=5,622). We conducted two telephone follow-up surveys with a representative subsample of 990 individuals in May and September 2020. The second panel (May) occurred when the government began implementing stay-at-home orders in many urban areas. We conducted the third panel in September when the government began relaxing some of the stricter control measures. The data allows us to compare individuals’ socioeconomic situation in informal settlements before the pandemic and at the beginning and the end of the period when strict regulations were implemented.

The article provides granular data about the changes in the socioeconomic situation of informal settlements during the pandemic. Our findings show that informal settlements are paying a high toll. Their employment loss is substantially larger than the loss reported in the general population, even compared to individuals with low educational levels. We also document that the immigrant population has been particularly affected during this health crisis. We complement this finding by describing the change in participation in government programs and the level of collaboration between neighbors that could potentially attenuate the consequences of employment loss among this particular group of the population. The pandemic triggered some degree of neighborhood collaboration, although the support neighbors provide to each other does not persist over time. The government has implemented special financial support programs (e.g., Bono COVID). However, despite the magnitude of the employment loss, those programs have not reached most informal settlers. Governments, the non-profit sector, and the community need to act immediately to provide economic support, urban infrastructure, and protections to individuals living in informal settlements. Governments need to innovate with policies targeted at the informal sector, immigrants, and single-headed households. This crisis may also be an opportunity for long-term improvements in these marginalized communities. The pandemic has worsened the deprivation of informal dwellers; without action, this may result in a long-term poverty trap for years to come.

The article proceeds as follows. Section 2 provides some contextual information on informal settlements in Chile. Section 3 describes the data and methods used. Section 4 presents the main findings of the data analysis. Section 5 discusses the findings and suggests some policy implications.

## Context: Informal Settlements in Chile

Relative to other Latin American countries, Chile has a small number of informal settlements. According to a cadaster conducted by the Ministerio de Vivienda y Urbanismo (44) (Ministry of Housing, MHU) in 2019, there are 802 informal settlements in Chile with 47,050 households. However, there is evidence that the number of settlements has rapidly increased in the pandemic [45]. The MHU defines these places as “precarious settlements of eight or more households that inhabit on a piece of land that they possess irregularly, without at least one of three basic services (electricity, water, and sewer), and where all the housing units composed a defined social and territorial unity”. The number of informal settlements has been growing in the last decade, from 657 in 2011 to 802 in 2019 [44]. Most of this increase has occurred in the country’s northern region, which is likely explained by an accelerated migration from neighboring countries in the last years. Many low-income individuals find a place to live in an informal settlement to transition to better opportunities in the country. 73% of all informal settlements in Chile are located in urban areas.

While the emergence of informal settlements in Chile’s urban areas dates back to the late nineteenth century, they became a relevant political and policy issue in the middle of the twentieth century when land invasions became politically salient [46-48]. People in those settlements organized to press the government for permanent housing solutions. Consecutive governments responded to the housing deficit through housing policies adopted or social programs designed for informal settlements [47,49]. In the 1960s and early 1970s, the public perceived informal settlements as politically organized communities that provided popular support for progressive social reforms. The democratically elected socialist government of Salvador Allende (1970-1973) had an ambiguous relationship with these communities, legitimizing many land invasions. Those years saw a substantial increase in the number of informal settlements formed in Chilean cities [47]. In a cadastre in 1972, the MHU counted 275 settlements, with approximately 450,000 individuals nationally [47]. This estimate suggests that, with half the Chilean population, three times more individuals lived in an informal settlement in 1972 than in 2019.

The military dictatorship (1973-1990) that followed Allende’s government took an aggressive approach toward informal settlements. The land invasions were particularly problematic for an administration that put a high value on private property to implement a market-oriented economic transformation. Thousands of families were forced to move to low-income, segregated housing projects located in the periphery of Chile’s urban areas [47]. A large-scale socioeconomic urban segregation pattern became the dominant reality of Chile’s urban development. This socioeconomic spatial segregation persists until today.

Since 1990, democratic governments have addressed housing deficits through a large-scale housing policy that, to a significant extent, has maintained the institutional architecture of the dictatorship [50,51]. A substantial portion of low and middle-income individuals have obtained access to formal housing through government programs, including many former residents of informal settlements. From a quantitative perspective, the success of the policy regime implemented since the 1990s in Chile is clear. However, a substantial failure of Chile’s housing policy has been the agglomeration of low-income individuals in segregated urban areas [52-54].

Chile has set up a policy scheme that has provided a substantial portion of informal settlers with a formal housing unit that they own [39]. However, the number of informal settlements has been growing in the past decade [49]. This growth, particularly in the north of Chile, may be partly explained by a growing influx of low-income migration from neighboring countries. Another reason relates to the dramatic increase in the price of urban land in many Chilean cities, which makes the government’s subsidies ineffective [49]. Housing quality and location have become increasingly important for families living in informal settlements [39]. Recent studies have suggested that for some essential dimensions of urban welfare, households living in informal settlements have access to a better geography of opportunities than those living in low-income housing projects where they own their dwellings [55,56]. These improved opportunities may reveal a mismatch between the government’s housing options for the low-income population and the actual expectations of those seeking access to formal housing, including those living in informal settlements.

Although living in an informal settlement involves tenure insecurity, insufficient access to basic services, and precarious housing conditions, for many residents these communities are perceived as a temporary stop as the family seeks a permanent, formal housing solution that would meet their social and economic demands. The COVID-19 pandemic can add another obstacle to the already complicated socioeconomic situation of the settlement dwellers.

## Data and Methods

We collected data in collaboration with TECHO-Chile (Techo), the Chilean branch of the largest Latin American non-governmental organization working with residents of informal settlements in the region. Our analysis is based on a three-wave panel (longitudinal) study of households living in Chile’s informal settlements. The baseline survey was conducted in person in August 2019 and obtained data from 5,622 household heads on different dimensions related to their life in the settlement.^i^

To examine the socioeconomic impact of the COVID-19 pandemic on informal dwellers, we conducted two telephone follow-up surveys with a representative random subsample of the total individuals interviewed at baseline. The first wave was conducted by phone in May and collected information on 990 households. The second wave was implemented in September and reached a total of 703 households from the first wave. Therefore, we built a panel of 703 individuals that were surveyed at three different moments over eleven months. Table 2 examines the balance across the three waves. The attrition rate was around 29% from wave two to wave three. However, based on the observable characteristics identified in 2019, the three waves are not significantly different in almost all observable categories (see supplementary material Table S2).

**Table 1.** Changes in employment status over time

|  | (1) | (2) | (3) |
| --- | --- | --- | --- |
| Post-1 | -0.249***<br>(0.0198) | -0.289***<br>(0.0284) | -0.347***<br>(0.0259) |
| Post-2 | -0.392***<br>(0.0188) | -0.405***<br>(0.0246) | -0.409***<br>(0.0220) |
| Constant | 0.732***<br>(0.00594) | 0.765***<br>(0.0164) | 0.782***<br>(0.0127) |
| Observations | 6,968 | 1,865 | 1,887 |
| R-squared | 0.079 | 0.214 | 0.253 |
| Individual FE | NO | NO | YES |
| Settlement FE | NO | YES | NO |
| Weight | NO | YES | YES |

**Notes** Coefficients are estimated under different model specifications. Columns (1) shows the result from a pooled cross-section regression, and coefficients Post-1 and Post-2 show the change in employment status for each particular period, relative to the pre-pandemic period measured in August 2019. Column (2) reproduces the same specification as column (1) but including settlement fixed effects and weights. Column (3) shows results, including individual fixed effects, as specified in equation (1) and include survey weights. Robust standard errors in parentheses. \* $p < 0.1$ , \*\* $p < 0.05$ , \*\*\* $p < 0.01$ .

**Table 2.** Changes in collaboration with neighbors over time

|  | (1) | (2) | (3) | (4) | (5) |
| --- | --- | --- | --- | --- | --- |
|  | Listens problems | Domestic Work | Money | Clothes and Food | Day Care |
| Post-1 | 0.0288<br>(0.0247) | 0.243***<br>(0.0253) | 0.00410<br>(0.0251) | 0.123***<br>(0.0236) | 0.169***<br>(0.0247) |
| Post-2 | 0.0481*<br>(0.0250) | 0.193***<br>(0.0258) | -0.0559**<br>(0.0243) | 0.0781***<br>(0.0248) | 0.0578**<br>(0.0250) |
| Constant | 0.640***<br>(0.0144) | 0.399***<br>(0.0149) | 0.358***<br>(0.0144) | 0.658***<br>(0.0143) | 0.326***<br>(0.0142) |
| Observations | 2,088 | 2,089 | 2,090 | 2,083 | 2,083 |
| R-squared | 0.003 | 0.073 | 0.006 | 0.021 | 0.035 |
| Individual FE | YES | YES | YES | YES | YES |
| Number of id | 703 | 703 | 703 | 703 | 703 |

**Notes.** The table shows coefficients for different outcomes as specified in each column. Each outcome variable is measured as an indicator variable for whether the respondent “*collaborates with his/her neighbors on*” issues identified by each column. Details about how each variable definition is available in Appendix Table A1. are estimated under different model specifications. All coefficients are estimated survey weights. Coefficients are estimated using equation (1), and all regressions include individual fixed effects. Robust standard errors in parentheses. \* $p < 0.1$ , \*\* $p < 0.05$ , \*\*\* $p < 0.01$ .

We aim to measure the changes in a set of outcomes over time. We exploit the individual-level panel data to analyze how each of the outcomes has changed over time. We estimate equation (1) as follows:

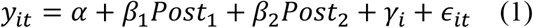

Where *β*_1_ and *β*_2_ are our coefficients of interest and represent the change in the outcome observed in May and September, 2020 respectively. In most specifications, we include an individual fixed effect (*γ*_*i*_) that controls for all time-invariant characteristics that may affect the different outcomes. We also discuss the robustness of the results to the inclusion of individual fixed effects. We analyze the evolution over time of outcomes related to employment status, cooperation among neighbors, and government assistance.

For employment status, we examine the magnitude of the results relative to other employment surveys. We carefully compare our results to different population groups from the ENE-Survey. ENE is the main employment survey run by Chile’s National Institute of Statistics, and we exploit individual microdata to examine the employment evolution during the same period.^ii^ We further examine the results for a particular subset of individuals available in the sample. In particular, we look at whether or not the results are driven by some underlying conditions related to informal dwellers that make them particularly vulnerable to the type of crisis triggered by the COVID-19 pandemic.

## Results

We present two sets of results that allow us to provide robust information about the socioeconomic impact of the COVID-19 health crisis on informal settlements. The first set refers to the observed changes in employment status over time, where we can examine their magnitude relative to other estimates obtained from national population surveys. Here we find a dramatic employment loss among informal dwellers during the pandemic, which is significantly larger than the labor decline in the general population. We complement this finding by examining changes in cooperative behavior among neighbors and the magnitude of the government policy take-up rate, which may ameliorate the economic impact suffered by individuals in the settlements. Although our information does not allow us to assess the overall change in economic wellbeing, we detect some responses oriented to reduce the dramatic loss in employment.

Table 1 summarizes the evolution of employment status. We compare coefficients under different model specifications. Columns (1) and (2) show results from a pooled cross-section model, while column (3) shows our preferred specification, including individual fixed effects. The results are remarkably robust across specifications. Post-1 and Post-2 capture the change in employment status observed in May and September 2020, respectively, compared to the pre-pandemic period in August 2019. After the pandemic, labor participation in the informal settlements drops 30 percentage points (Table 1, column 3, Post-1; p<0.01), representing a 50 percent drop relative to the pre-pandemic level. The decrease in September is even more considerable. Compared to August 2019, labor force participation decreases by around 40 percentage points (Table 1, column 3, Post-2, p<0.01), from a pre-pandemic level of 75 percent.

To examine the magnitude of the decrease in employment, we compare our results with a nationally representative survey. Figure 1 compares the evolution of labor participation over time for different groups: national population, individuals with incomplete formal education (less than secondary) residing in the same municipalities as our sample, and informal dwellers. We show these results in two figures. Fig. 1A examines the evolution of the percentage of people employed at any given moment, and Fig. 1B shows the change relative to the observed levels for each category in the pre-pandemic period. Relative to the national population, informal dwellers usually exhibit a higher level of participation in the labor market [57].

**Figure 1:**
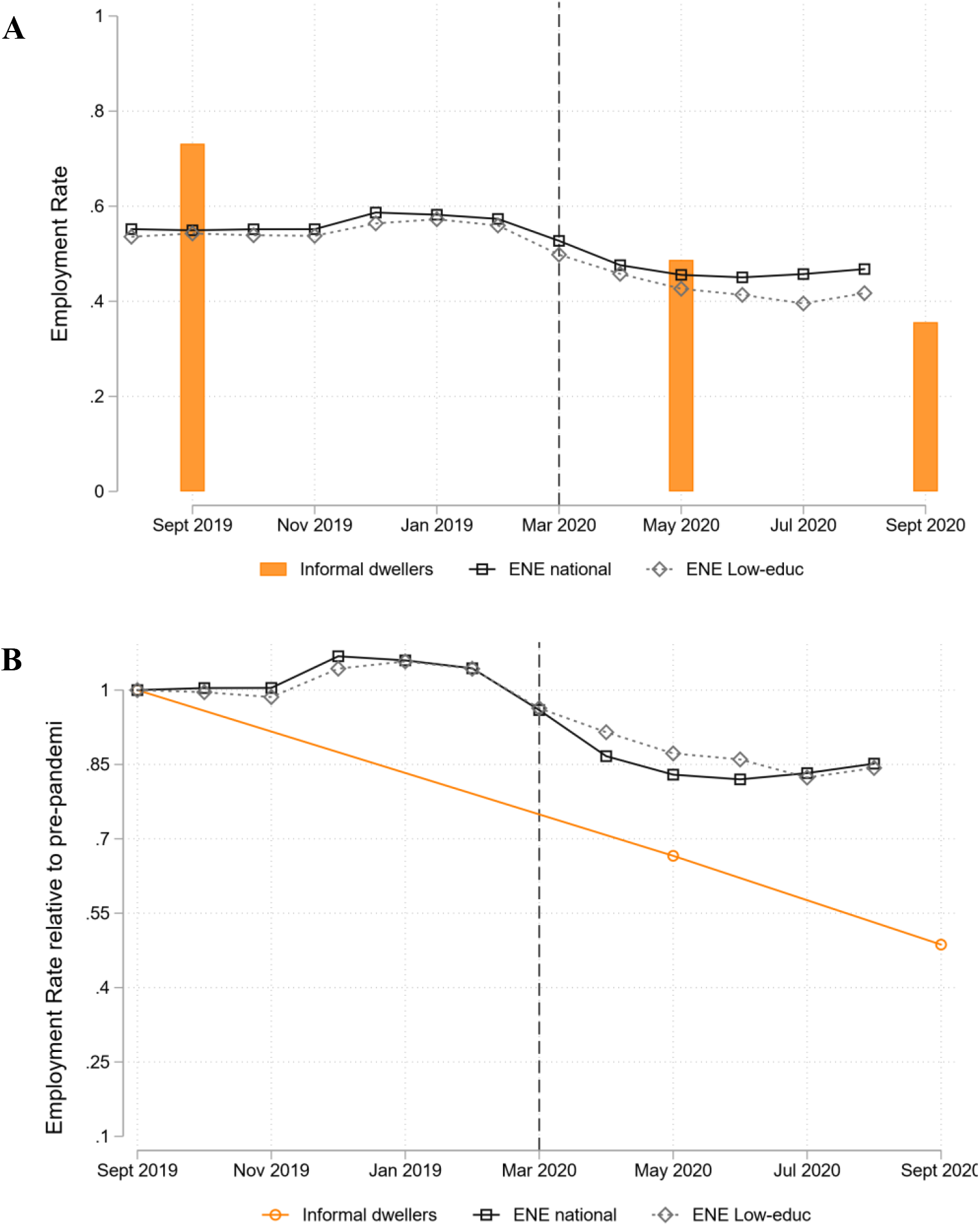
Evolution of the employment rate as the proportion of individuals employed (A) and relative to pre-pandemic levels (B). The figure shows the evolution of the employment rate before and after the COVID-19 pandemic period for different survey series. Panel A shows the employment rate as the proportion of people employed divided by the population older than fifteen years old on each survey. Panel B shows the employment rate evolution relative to the average level observed during the pre-pandemic months observed on each series. The vertical dashed line marks the beginning of the pandemic in Chile. From ENE monthly surveys, we plotted both the evolutions of the nationally-representative sample and a subsample of respondents who reported less than complete secondary education and were residing in municipalities with at least one informal settlement. Information on informal dwellers was obtained from Techo-Panel Survey 2019-2020.

Figure 1 shows that relative to the national population, employment loss is substantially larger among informal dwellers. The decrease in employment status is even larger than for individuals with lower education levels living in the same municipalities. Comparing the evolution of individuals in the same municipalities may alleviate concerns about localized lockdowns implemented at the municipality-level. These lockdowns may affect people differently, depending on the district where they live [6]. Although one can imagine several reasons why these two populations may differ, such as the type of employment, the magnitude of the difference shows that unemployment affected informal dwellers more severely.

We further examine the extent to which the larger drop in employment can be explained with some observable attributes of informal dwellers that have been more affected by the pandemic at the national level. Figure 2A examines whether the large proportion of immigrants living in informal settlements may account for the larger employment loss. Figure 2B shows the employment impact disaggregated by gender.

**Figure 2:**
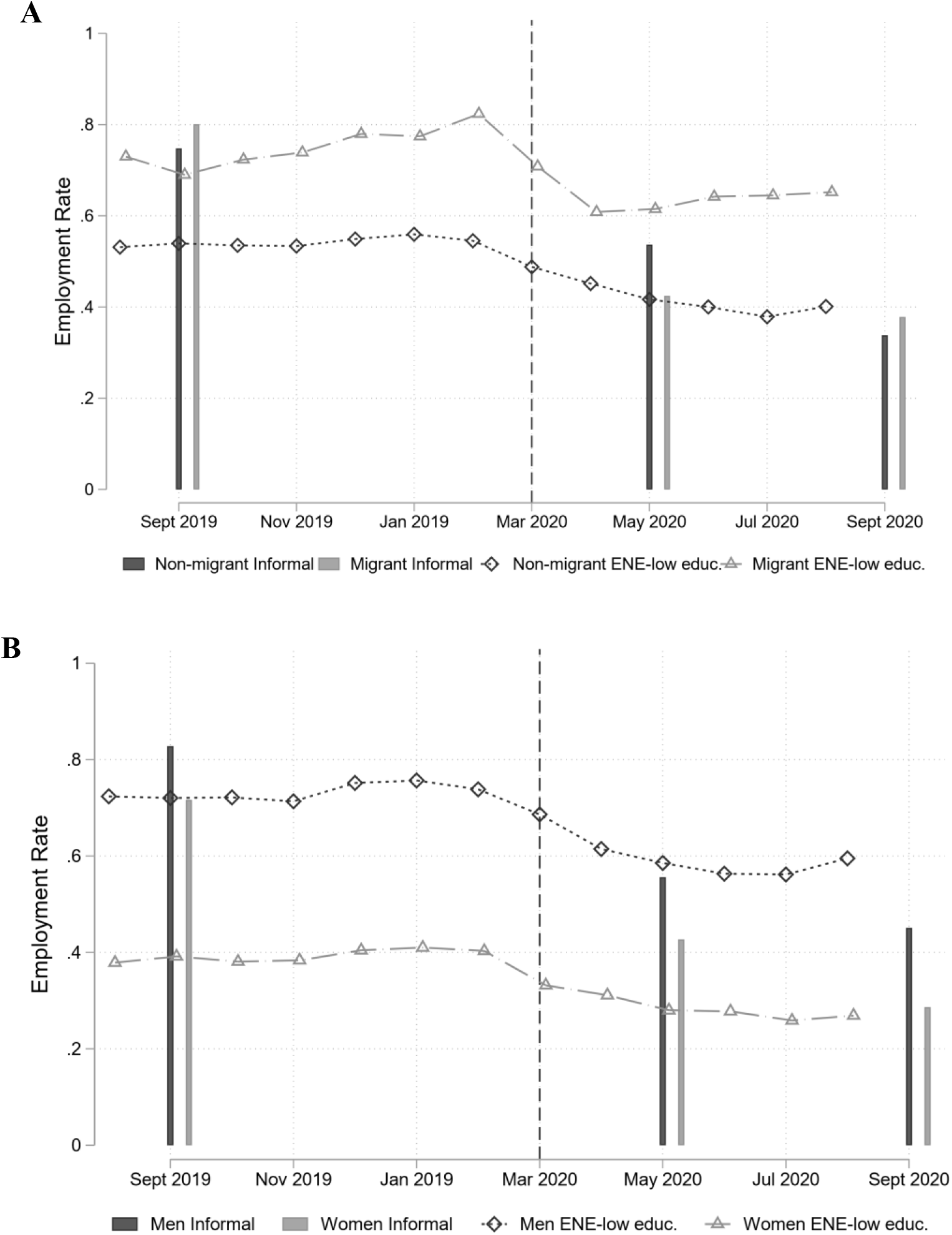
Employment rate evolution, by immigration status (A) and gender (B). Evolution of employment rate before and after the COVID-19 pandemic period for different survey series. The employment rate is measured as the proportion of people employed divided by the population older than fifteen years old on each survey. Panel A splits each sample by the respondent’s immigration status, and Panel B splits each sample by respondent’s gender. The vertical dashed line marks the beginning of the pandemic in Chile. ENE monthly surveys show the evolutions of a subsample of respondents who reported less than complete secondary education and resided in municipalities with at least one informal settlement. Information on informal dwellers was obtained from Techo-Panel Survey 2019-2020.

Figure 2 shows that employment loss in May was particularly large among immigrants. Immigrants in both surveys display a higher employment rate before the pandemic period and a dramatic loss at the beginning of the pandemic. Among immigrant respondents with incomplete secondary education from the representative national population survey, the employment rate drops from 80 to 60 percent between February and April. Informal immigrant dwellers suffered a larger drop, from 80 to 40 percent between September 2019 and May 2020.^iii^

Interestingly, after April, the immigrant population from the national population survey exhibits a growing trend in employment up to 65 percent in August. This contrasts with the evolution of the Chilean respondents where employment rate drops from 55 percent in February, to 45 percent in April, and continues falling to 40 percent in August. Among informal dwellers, we observe a similar trend across groups with an important difference in scale. Relative to Chilean settlement dwellers, employment loss among immigrants was 15 percent larger in May. In September, the employment rate decreased further for both groups up to 40 percent with no significant difference across groups.^iv^

We now analyze the evolution of collaboration among neighbors, a dimension of informal settlements’ social capital, which may mitigate some of the economic hardships we have described for the informal settlements’ population. The questions analyzed refer to whether respondents provide help to their neighbors in their communities. Table 2 shows the results. All Post-2 coefficients in Table 2 suggest a large change in social collaboration among neighbors. Although for “listening to problems” and “lending money” we detect no significant change in May, the variation is significant in the September wave. In the case of “listening to problems” the increase over time may reflect an increase in social interactions. This increase may not necessarily have shown up at the beginning of the mandatory lockdown when people complied more strictly with social distancing measures. However, that strict interpretation should be viewed with caution since we detect a deterioration in the percentage of respondents who report helping neighbors with domestic work, clothes and food, and children care (Table 2, columns 2, 4, and 5 respectively). The pandemic’s beginning may have triggered a large increase that persists over time with a slight intensity decrease.

We finally analyze the take-up evolution of government programs implemented to reduce the consequences of employment loss during this period. In May and September 2020, we incorporated questions regarding participation in the unemployment insurance program and the COVID cash transfer program (“Bono COVID”) that the government put in place in response to the pandemic crisis. Through Bono COVID, low-income households can receive a cash benefit of approximately US $70.

Table 3 shows that participation in the unemployment insurance program among informal dwellers increased from 6 to 17 percent between May and September, 2020. An essential limitation of the unemployment insurance program’s ability to ameliorate the employment loss is the high level of informal employment, leaving a large group of informal dwellers ineligible for this program. In fact, we observe that roughly a quarter of the respondents that lost employment between 2019 and 2020 claimed the unemployment insurance benefit. Similar to the unemployment insurance program, participation in Bono COVID program increased from 33 to 37 percent during 2020.^v^ Although take-up ratio for this program was higher among those who never reported any employment (56 percent) or those who lost employment after the pandemic (53 percent), more than half of the respondents reported having not received this benefit during the pandemic period.

**Table 3.** Changes in programs participation between May and September, 2020.

|  | (1) | (2) | (3) | (4) | (5) | (6) |
| --- | --- | --- | --- | --- | --- | --- |
|  | Unemp.<br>Insurance | Unemp.<br>Insurance | Unemp.<br>Insurance | Covid<br>transfer | Covid<br>transfer | Covid<br>transfer |
| Post-2 | 0.112***<br>(0.0175) | 0.110***<br>(0.0175) | 0.114***<br>(0.0161) | 0.0431<br>(0.0264) | 0.0418*<br>(0.0245) | 0.0400**<br>(0.0163) |
| Constant | 0.0606***<br>(0.00937) | 0.0611***<br>(0.00984) | 0.0594***<br>(0.00816) | 0.327***<br>(0.0184) | 0.329***<br>(0.0170) | 0.329***<br>(0.00825) |
| Observations | 1,381 | 1,363 | 1,381 | 1,387 | 1,369 | 1,387 |
| R-squared | 0.030 | 0.117 | 0.074 | 0.002 | 0.211 | 0.009 |
| Individual FE | NO | NO | YES | NO | NO | YES |
| Settlement FE | NO | YES | NO | NO | YES | NO |

**Notes.** The table shows coefficients for different outcomes as specified in each column. Each outcome variable is measured as an indicator variable for whether the respondent “*receives resources from*” sources identified by each column. Details about how each variable definition is available in Appendix Table A1. All coefficients are estimated using survey weights. Coefficients are estimated using equation (1). Robust standard errors in parentheses. \* $p < 0.1$ , \*\* $p < 0.05$ , \*\*\* $p < 0.01$ .

## Discussion & Policy Implications

The COVID-19 pandemic has altered everyday life dramatically for most of the world’s population, imposing new costs, restrictions, and risks on everyday life. These changes also affect individuals living in informal settlements, but their options to cope with these changes are worse. This study’s empirical evidence of the socioeconomic effects of the pandemic in informal settlements shows a grim situation. Compared to the baseline in August 2019, our results showed a dramatic drop in employment status in May and September 2020. This drop was more extensive for settlement dwellers than for the rest of the population and also when compared to individuals with low education (a proxy for low income) in the same districts. Our analysis shows that the pandemic has triggered some level of neighborhood cooperation on some basic needs, but that cooperation has decreased over time. Last, we describe the number of informal households that have received assistance from the government, which has increased during the pandemic but has not reached the majority of the population examined.

There is substantial evidence of the harmful effects of the pandemic and viral transmission control strategies, such as lockdowns and school closures, on economic activity. These effects have resulted in substantial employment loss [7,9-11,58]. Residents of informal settlements are more economically vulnerable to external shocks in the economy than the general population because a large proportion perform informal jobs earning subsistence wages. Our findings are consistent with this evidence.

The pandemic has also exacerbated pre-existing social inequalities. For example, our results also show that the drop in economic activity has more negatively affected immigrants than non-immigrant residents of informal settlements. The situation of the most vulnerable populations is maybe even worse than we found in our study, as there is increasing evidence of xenophobia, racism, and stigma against the poor and migrants during the COVID-19 pandemic [59-62].

Our results also suggest neighborhood cooperation within informal settlements increased at the onset of the pandemic but has decreased over time. This decrease may be an essential source of vulnerability, as individuals living in these settlements often depend on local networks to access resources, including food, labor, and childcare [26]. We also found that immigrant householdsare are substantially less likely to receive any government assistance, such as the Bono COVID or in-kind food transfers. Informal settlements often attract less public assistance because they are installed on land without a proper legal title [34]. This complex relationship with governments could be more problematic during the pandemic. Communities sometimes cope with the lack of aid from government offices by developing strong social networks among neighbors and also with the aid of NGOs and other private institutions.

Strategies to prevent the spread of the virus need to consider the chronic vulnerabilities of individuals living in informal settlements and the comparative advantages of these communities. Some of these communities’ vulnerabilities to infectious diseases include inadequate water, sewage, and hygiene infrastructure; insufficient or nonexistant waste collection; material deprivation; space limitations; and overcrowding [26,63,64]. These characteristics need to be considered when implementing large scale regulations and policies to control a pandemic, as the livelihoods of people in informal settlements are more vulnerable to external shocks in the economy. Some researchers have conceptualized these communities as “poverty traps”, which prevent low-income families from accumulating the economic and social resources necessary to improve their disadvantaged situation [37]. Other authors have provided a more complex picture of informal settlements, conceiving these urban enclaves as low-cost housing options for vulnerable individuals that aim to take advantage of the access that the settlements provide to the economic and social opportunities that a city brings [36]. Individuals in some informal urban settlements may have better access to economic and social urban opportunities – such as connectivity, jobs, social networks, and community organizations – than low-income individuals in government-subsidized housing projects [39,55,56]. Urban informal settlements may be a relevant source of economic activity and income in a city and may run social, political, and cultural initiatives [39,65,66].

Governments, non-profits, and the community need to act quickly to provide economic support, urban infrastructure, and protections to individuals living in informal settlements. The value of lives saved and disease burden averted will probably far exceed the economic costs of providing immediate support to families facing severe hardship [20]. Schools provide essential meals and nutrition to children living in poverty, including those in informal settlements. The closure of schools, implemented nationally in Chile since March 16 [40], likely has affected child nutrition and other essential services, such as routine vaccination programs [67]. This urgent support should not come at the expense of long-term efforts to improve informal settlements and their residents’ other immediate needs [21]. Governments need to innovate with policies targeted at the informal sector, including immigrants and single-headed households. This crisis may also be an opportunity for long-term improvements in these marginalized communities.

We have provided quantitative evidence of the effects of the COVID-19 pandemic and mitigation strategies in informal urban settlements. The emergence and persistence of informal settlements in urban areas represent a complex policy issue, as these settlements may offer advantages compared to government-provided-low-income housing. Strategies to prevent the spread of the virus need to consider the chronic vulnerabilities of individuals living in informal settlements and the comparative advantages of these communities. Informal settlements are densely populated areas where compliance with social distancing is challenging. This crisis has expanded the spatial inequalities of urban areas, affecting the population living in informal settlements, many of whom may end up in a long-term poverty trap for years to come.

## Data Availability

The data used for this study are available from the corresponding author upon reasonable request.

## Acknowledgments

We thank Laura Marshall (P. Universidad Católica de Chile) for outstanding research assistance with data analysis. We also want to thank the excellent work of Catalina Rufs (P. Universidad Católica de Chile) with the data collection and data analysis for the project of which this paper is part of. We thank the staff of TECHO-Chile and all their volunteers who work in the implementation of the survey and the staff of the Dirección de Estudios Sociológicos UC for technical support.

## Ethics approval and consent to participate

The study was approved by the Comité Ético Científico de Ciencias Sociales, Artes, y Humanidades of the Pontificia Universidad Católica de Chile (Protocol ID: 200413004). We obtained participant’s informed consent by telephone, to avoid the risk of CoVID-19 contagion.

## Supplementary Material

**Table S1.** Techo survey households by region of residence, and relative to national census of informal settlements

| REGION | MINVU 2019 | Percentage | TECHO | Percentage |
| --- | --- | --- | --- | --- |
| Tarapacá | 4,084 | 9% | 864 | 15% |
| Antofagasta | 7,641 | 16% | 937 | 17% |
| Atacama | 4,648 | 10% | 556 | 10% |
| Coquimbo | 1,019 | 2% | 343 | 6% |
| Valparaíso | 11,228 | 24% | 710 | 13% |
| OHiggins | 1,271 | 3% | 0 | 0% |
| Maule | 152 | 0% | 0 | 0% |
| Biobío y Ñuble | 6,747 | 14% | 543 | 10% |
| La Araucanía | 351 | 1% | 0 | 0% |
| Los Lagos | 2,090 | 4% | 155 | 3% |
| Aysén | 252 | 1% | 0 | 0% |
| Magallanes | 53 | 0% | 0 | 0% |
| Metropolitana | 5,991 | 13% | 1,378 | 25% |
| Los Ríos | 1,197 | 3% | 121 | 2% |
| Arica y Parinacota | 326 | 1% | 0 | 0% |
| Total | 47,050 |  | 5,607 |  |

**Notes.** Table describes the region of residence of the TECHO and MINVU survey. In 2019, the Chilean Ministry of Housing and Urbanism surveyed all informal settlements in the country, and reported a total of 47,050 households.

**Table S2:** Balance across survey waves.

|  | All 2019<br>respondents | Wave 1 & 2019<br>respondents | p-value<sup>†</sup> | Waves 1-2 &<br>2019 respondents | p-value<sup>‡</sup> |
| --- | --- | --- | --- | --- | --- |
| Employment | 0.7315<br>(0.4432) | 0.7315<br>(0.4434) | 0.9985 | 0.7467<br>(0.4352) | 0.3901 |
| Migrant | 0.3851<br>(0.4867) | 0.3983<br>(0.4898) | 0.435 | 0.3957<br>(0.4893) | 0.5913 |
| Persons in household | 3.4024<br>(3.4108) | 3.3292<br>(1.6916) | 0.3024 | 3.3232<br>(1.6922) | 0.3158 |
| Children in household | 1.2000<br>(1.2257) | 1.1624<br>(1.1602) | 0.3552 | 1.1806<br>(1.174) | 0.6844 |
| Persons >65 years in household | 0.0982<br>(0.3931) | 0.0915<br>(0.3497) | 0.5867 | 0.1026<br>(0.376) | 0.7726 |
| Persons in household with<br>chronic diseases | 0.3531<br>(0.6071) | 0.3998<br>(0.6363) | 0.0334 | 0.4263<br>(0.6514) | 0.0051 |
| Adults who did not finish school<br>in household | 0.6072<br>(0.4884) | 0.5992<br>(0.4903) | 0.6373 | 0.5910<br>(0.492) | 0.4136 |
| Arrived to informal settlement<br>because of economic reasons | 0.6440<br>(0.4789) | 0.6449<br>(0.4788) | 0.957 | 0.6443<br>(0.4791) | 0.9853 |
| Previously lived in an informal<br>settlement | 0.1023<br>(0.3031) | 0.0904<br>(0.287) | 0.2383 | 0.0954<br>(0.2939) | 0.5603 |
| Arrived to informal settlement<br>more than three years ago | 0.4598<br>(0.4984) | 0.4844<br>(0.500) | 0.158 | 0.4985<br>(0.5004) | 0.056 |
| Poor quality walls | 0.4298<br>(0.4951) | 0.4361<br>(0.4962) | 0.7164 | 0.4311<br>(0.4956) | 0.9472 |
| Poor quality ceiling | 0.3998<br>(0.4899) | 0.3933<br>(0.4887) | 0.7046 | 0.3968<br>(0.4896) | 0.8793 |
| Poor quality floor | 0.4501<br>(0.4975) | 0.4329<br>(0.4957) | 0.3264 | 0.4169<br>(0.4934) | 0.1003 |
| Have access to drinking water | 0.7923<br>(0.4057) | 0.8022<br>(0.3986) | 0.4872 | 0.8066<br>(0.3952) | 0.379 |
| Have trust in neighbors | 0.5465<br>(0.4979) | 0.5537<br>(0.4974) | 0.6781 | 0.5664<br>(0.4959) | 0.3212 |
| Observations | 5,494 | 955 |  | 682 |  |

**Notes** <sup>†</sup>*p-values* for All- Wave1. <sup>‡</sup>*p-values* for All- Waves1,2. The table shows the average values for each variable and sample in the 2019 survey. Standard deviations are in parentheses. We calculate the *p-value* of the difference across sample surveys for a different combination of samples identified in each column. The original 2019 survey was conducted in person in August 2019. The first and second waves correspond to a random sample of the original respondents re-surveyed in May 2020 and September 2020.

**Figure S1.**
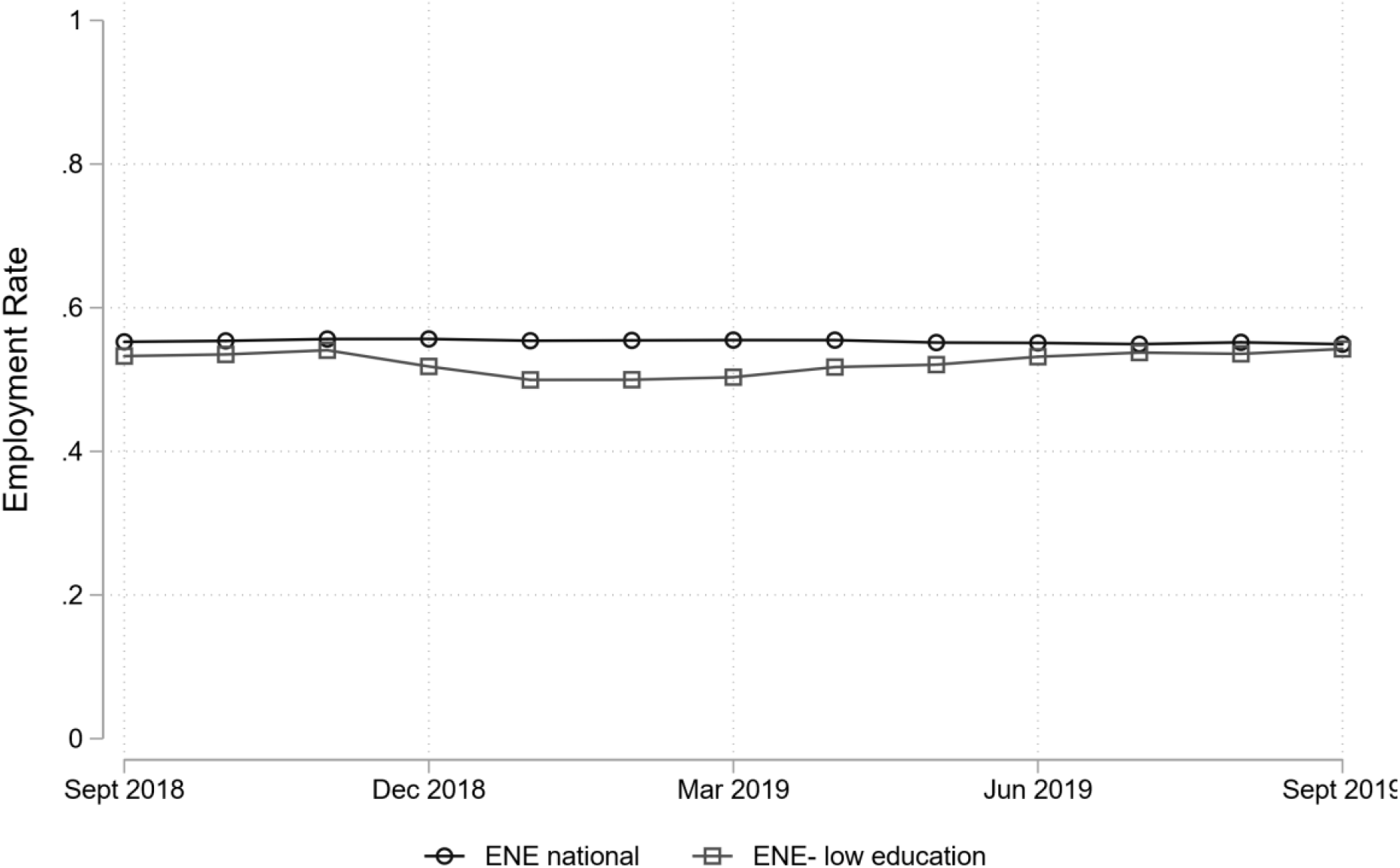
Employment rate evolution during 2018-2019. Evolution of employment rate during 2018-2019 following similar months described in figures 1 and 2 by immigration respondent status before and after the COVID-19 pandemic period for different survey series. The figure shows the employment rate as the proportion of people employed divided by the population older than fifteen years old on each survey. From ENE monthly surveys, we plotted a subsample of respondents who reported less than complete secondary education and were residing in municipalities with at least one informal settlement.

**Table S3.**
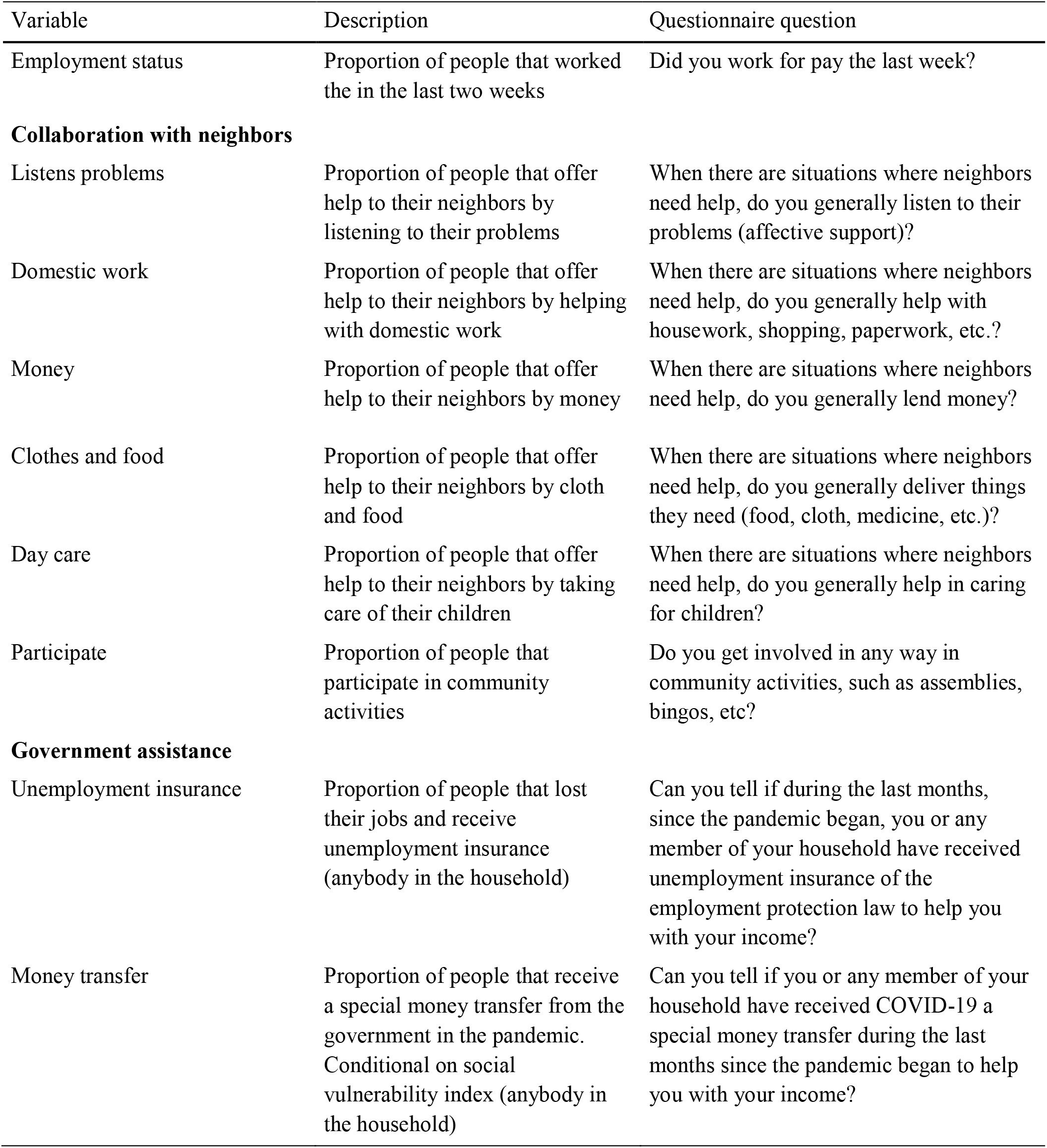
Variable definition

**Table S4.** Heterogenous change in employment status

|  | (1) | (2) | (3) | (4) | (5) | (6) |
| --- | --- | --- | --- | --- | --- | --- |
| Post-1 | -0.211***<br>(0.0383) | -0.221***<br>(0.0377) | -0.286***<br>(0.0346) | -0.272***<br>(0.0430) | -0.272***<br>(0.0433) | -0.310***<br>(0.0407) |
| Post-2 | -0.410***<br>(0.0331) | -0.407***<br>(0.0325) | -0.407***<br>(0.0291) | -0.377***<br>(0.0393) | -0.372***<br>(0.0391) | -0.377***<br>(0.0356) |
| Immigrant | 0.0530<br>(0.0334) | 0.0149<br>(0.0415) |  |  |  |  |
| Post-1x Immigrant | -0.165***<br>(0.0581) | -0.167***<br>(0.0581) | -0.148***<br>(0.0523) |  |  |  |
| Post-2x Immigrant | -0.0129<br>(0.0516) | -0.0121<br>(0.0508) | -0.0215<br>(0.0448) |  |  |  |
| Female |  |  |  | -0.111***<br>(0.0324) | -0.0878**<br>(0.0342) |  |
| Post-1x Female |  |  |  | -0.0179<br>(0.0569) | -0.0331<br>(0.0570) | -0.0661<br>(0.0526) |
| Post-2 x Female |  |  |  | -0.0533<br>(0.0504) | -0.0574<br>(0.0499) | -0.0554<br>(0.0450) |
| Constant | 0.748***<br>(0.0219) | 0.765***<br>(0.0242) | 0.789***<br>(0.0128) | 0.828***<br>(0.0236) | 0.815***<br>(0.0247) | 0.782***<br>(0.0127) |
| Observations | 1,807 | 1,785 | 1,807 | 1,887 | 1,865 | 1,887 |
| R-squared | 0.137 | 0.225 | 0.269 | 0.146 | 0.226 | 0.254 |
| Individual FE | NO | NO | YES | NO | NO | YES |
| Slum FE | NO | YES | NO | NO | YES | NO |

**Notes:** Coefficients are estimated under different model specifications. Columns (1) and (4) show results from a pooled cross-section regression, and coefficients Post-1 and Post-2 show the change in employment status for each particular period, relative to the pre-pandemic period measured in August 2019. Column (2) and (5) reproduce the same specification as column (1) and (4) but including slum fixed effects. Column (3) and (6) show results, including individual fixed effects as specified in equation (1). All coefficients are estimated using survey weights. Robust standard errors in parentheses. \* $p < 0.1$ , \*\* $p < 0.05$ , \*\*\* $p < 0.01$ .

**Table S5.** Heterogenous change in program participation

|  | (1) | (2) | (3) | (4) | (5) | (6) | (7) | (8) |
| --- | --- | --- | --- | --- | --- | --- | --- | --- |
|  | Unemp.<br>Insurance | Unemp.<br>Insurance | Covid<br>transfer | Covid<br>transfer | Unemp.<br>Insurance | Unemp.<br>Insurance | Covid<br>transfer | Covid<br>transfer |
| Post-2 | 0.0873***<br>(0.0208) | 0.0895***<br>(0.0192) | 0.0309<br>(0.0341) | 0.0332<br>(0.0239) | 0.109***<br>(0.0279) | 0.114***<br>(0.0262) | 0.0874**<br>(0.0362) | 0.0805***<br>(0.0277) |
| Immigrant | -0.000560<br>(0.0283) |  | -0.192***<br>(0.0453) |  |  |  |  |  |
| Post-2 x Immigrant | 0.0525<br>(0.0373) | 0.0570*<br>(0.0339) | 0.0151<br>(0.0492) | 0.00564<br>(0.0323) |  |  |  |  |
| Female |  |  |  |  | 0.0133<br>(0.0220) |  | 0.157***<br>(0.0352) |  |
| Post-2 x Female |  |  |  |  | 0.00124<br>(0.0357) | 0.00100<br>(0.0332) | -0.0780<br>(0.0486) | -0.0696**<br>(0.0340) |
| Constant | 0.0606***<br>(0.0151) | 0.0588***<br>(0.00821) | 0.413***<br>(0.0274) | 0.330***<br>(0.00831) | 0.0534***<br>(0.0164) | 0.0594***<br>(0.00817) | 0.239***<br>(0.0253) | 0.329***<br>(0.00822) |
| Observations | 1,310 | 1,328 | 1,313 | 1,331 | 1,363 | 1,381 | 1,369 | 1,387 |
| R-squared | 0.119 | 0.079 | 0.229 | 0.008 | 0.117 | 0.074 | 0.225 | 0.016 |
| Individual FE | NO | YES | NO | YES | NO | YES | NO | YES |
| Slum FE | YES | NO | YES | NO | YES | NO | YES | NO |

**Notes:** Coefficients are estimated under different model specifications following equation (1), and different combinations of fixed effects identified at the bottom of each column. Post-2 represents the change in the outcome in September 2020, relative to the observed level in May 2020. All coefficients are estimated using survey weights. Robust standard errors in parentheses. \* $p < 0.1$ , \*\* $p < 0.05$ , \*\*\* $p < 0.01$ .

## Endnotes

i See supplementary material Table S1 that describes the location of individuals surveyed in 2019, relative to the sample of slums identified in each Chilean region, according to government official records.

ii Encuesta Nacional de Empleo (ENE Survey) is a monthly national survey that surveys a random sample of the Chilean household. The survey contains information of all individuals older than 15 years within a selected household, and it corresponds to the main instrument to monitor the evolution of employment, among other labor market outcomes. The sample of the survey considers approximately 12.000 households which correspond to approximately 35,000 individuals each month. The survey considers as employees those who answer with “yes” to the question if last week they worked for at least one hour. This survey is managed by the Chilean national Institute of Statistics (Instituto Nacional de Estadísticas, INE).

iii While we do not observe monthly evolution of employment among informal dwellers, an obvious concern may relate to employment seasonality. Given the magnitude of the drop, seasonality seems to be a second order issue that could affect the magnitude of the observed change. By using the same nationally representative survey during 2019, we observe a fair degree of stability over time among the national population and the subgroup of respondent with incomplete education residing in municipalities with at least one slum. See supplementary material Figure S1.

iv In supplementary material Table S4 we analyze these results using our individual-level panel data and including heterogenous changes by immigrant status and gender. Coefficient results show a disproportionate drop among informal immigrant dwellers in May, but no significant difference between Chileans and immigrants in September. In terms of gender, we observe a larger drop -- but not statistically significant at conventional levels -- among females.

v We analyze heterogenous responses by immigration status and gender in supplementary material Table S5. In this case, we are constrained by the fact that these questions were later incorporated in the 2020 surveys. We observe several important facts. First, while we do not detect differences between Chileans and immigrants in any of the two periods for the unemployment insurance program, differences in COVID-transfers are substantial. In May, 2020, while 40 percent of Chilean informal dwellers had received the transfer, only 20 percent of immigrants had participated. In terms of gender, we observe that women exhibit a 16 percent higher participation rate in COVID-transfer program in May, 2020 and the gender gap drops by 7 percentage points in September, 2020. In the case of unemployment insurance program participation, we observe no difference by gender among informal dwellers in any of the two periods of analysis.

